# Optimizing Aqueous Humor Liquid Biopsy: Safety and Performance of a Short, Low-Dead-Space Ophthalmic Needle for Anterior Chamber Paracentesis

**DOI:** 10.64898/2026.08.26.26361364

**Authors:** Aneal M. Singh, Tsai-Chu Yeh, Charles DeBoer, Ahmad Al-Moujahed, Jonathan B. Lin, Stephen A. Smith, Steven Sanislo, Kholood Janjua, Tai-Chi Lin, David R. P. Almeida, Prithvi Mruthyunjaya, Vinit B. Mahajan

**Author notes:** Co-first authors (Aneal M. Singh, Tsai-Chu Yeh, Vinit B. Mahajan). **Correspondence:** Vinit B. Mahajan, M.D., Ph.D., Molecular Surgery Laboratory, Byers Eye Institute, Department of Ophthalmology, Stanford University, Palo Alto, CA 94304, USA.

## Abstract

**Purpose:** To evaluate the safety, procedural performance, sample recovery, and surgeon preference of an ophthalmic needle designed specifically for anterior chamber (AC) paracentesis.

**Methods:** In this multicenter study, AC paracentesis was performed in clinic and operating-room settings using a 32-gauge*×* 4-mm needle with low dead space. The procedure was evaluated using a standardized physician survey. Prespecified outcomes included procedure-related adverse events (primary outcome), needle entry and handling, aspiration and sample recovery, comparative performance versus a 30-gauge needle, and physician preference for future use.

**Results:** A total of 110 needle uses by eight surgeons were included. No ocular complications occurred, including lens or iris injury, hyphema, AC collapse, wound leak, hypotony, infection, or retinal complication, and no procedure required needle exchange or conversion to another device. Two technical events without ocular sequelae were noted, in which needle entry was partial thickness and did not reach the AC (1.8%; exact 95% CI, 0.2%–6.4%). Physicians rated needle entry, handling, and sample recovery as good or excellent. Compared with a 30-gauge needle, the study needle was rated as at least comparable across all assessed domains. All surgeons rated it better or much better for intra-procedural safety and preferred it for future AC taps.

**Conclusions and Relevance:** This short, 32-gauge low-dead-space ophthalmic needle demonstrated a favorable safety profile and was preferred over a 30-gauge needle by all surgeons. As aqueous humor liquid biopsy expands in clinical diagnostics and trials, an ophthalmic-specific needle design may help improve the consistency and safety of aqueous humor collection for molecular analysis and broader clinical use.

## Introduction

Aqueous humor (AH) liquid biopsy has become an important tool in translational ophthalmic research, enabling molecular characterization of ocular disease in living patients.^1^ Anterior chamber (AC) paracentesis itself is a long-established procedure, traditionally performed to acutely lower intraocular pressure and to diagnose intraocular inflammation and infection, and now increasingly used in ocular oncology. Aqueous polymerase chain reaction (PCR) testing informs the workup of infectious uveitis and endophthalmitis,^2,3^ and tumor-derived DNA and liquid-biopsy proteomics have shown genomic alterations and metastatic risk in uveal melanoma.^4–6^ In vitreoretinal lymphoma, detection of the MYD88 L265P mutation now offers a less invasive route to diagnosis and longitudinal assessment.^7^ More recently, applications have expanded beyond oncology to common ocular diseases through proteomic, metabolomic, and multi-omic profiling.^8^ Aqueous biomarkers have been used to evaluate intraocular pharmacokinetics in clinical trials^9,10^ and to predict individual treatment response and characterize disease mechanisms across a widening range of retinal diseases, including diabetic retinopathy and age-related macular degeneration.^8,11–14^ As AH liquid biopsy extends from specialized practice into these prevalent conditions, a safe, standardized, and surgeon-trusted collection procedure becomes central to expanding its accessibility in routine clinical care.

The number and complexity of molecular applications are growing in ophthalmology and the quality and efficiency of specimen acquisition matter more than ever. Standardized, low-volume collection is essential to minimize sample loss and preserve material for downstream analysis. AC paracentesis provides direct access to AH, yet the procedure is commonly performed with needles repurposed from general hypodermic use rather than devices designed for low-volume ophthalmic sampling. These general-purpose needles have shaft lengths exceeding those required for clear-corneal entry, resulting in unnecessary intraocular needle exposure and increasing the potential for inadvertent contact with intraocular structures; lens contact, which may result in traumatic cataract, has been reported following AC paracentesis.^15^ Conventional hub dead space may further retain a meaningful proportion of a 50-to 100-µL specimen, and opaque or colored hubs can make it difficult to see clear aqueous fluid entering the syringe.^1,16–18^ This is a critical issue as splitting the small volume fluid for multi-omic or drug pharmacokinetic testing becomes a growing need.^8^ Needle design therefore shapes both procedural control and specimen recovery. The 32-gauge needle evaluated in this study was designed around exactly these constraints: a 4-mm shaft that limits how far the needle can advance into the eye, and a transparent, low-dead-space hub that reduces retained volume and allows physicians to watch fluid enter during aspiration.

The safety of AC paracentesis itself is increasingly established. A retrospective electronic health record analysis of 1,418 aqueous biopsies obtained during intraocular surgery identified a single minor complication and no cases of endophthalmitis,^19^ and a recent retrospective multicenter study of 1,203 paracentesis procedures in pediatric patients with retinoblastoma, performed in small eyes with shallow anterior chambers, reported a similarly favorable safety profile.^20^ Around the world, however, AC taps are still performed with a heterogeneous assortment of repurposed needles.^21^ Existing safety data also derive predominantly from retrospective chart review. In this multicenter study spanning academic, community, and international practice settings, we prospectively evaluated the safety and surgeon-reported performance of a short 32-gauge low-dead-space ophthalmic needle. Standardizing a safe, reproducible sampling technique will be key to expanding AH liquid biopsy from specialized centers to broader molecular applications and, ultimately, routine clinical care.

## Materials and Methods

### Study Design and Ethical Approvals

This multicenter study evaluated clinical use of a 32-gauge ophthalmic needle for AC paracentesis at the Byers Eye Institute at Stanford University (Palo Alto, California), Erie Retina Research and Erie Retinal Surgery (Erie, Pennsylvania), and Taipei Veterans General Hospital (Taipei, Taiwan). Participating surgeons completed a cross-sectional device assessment survey after accumulating clinical experience with the device. The protocol was approved by the respective Institutional Review Boards, and the study adhered to the tenets of the Declaration of Helsinki and, at United States sites, the Health Insurance Portability and Accountability Act. Written informed consent or an approved waiver of consent was obtained in accordance with the study protocol.

### Patient and Surgeon Cohorts

Eligible procedures were diagnostic, therapeutic, or research-associated AC paracentesis performed in adults undergoing ophthalmic care. All procedures were performed by ophthalmologists with prior experience performing AC taps. For each procedure, we documented the clinical service (ocular oncology, general retina, or genetic retinal disease) and setting (outpatient clinic or operating room). Procedure and physician characteristics are summarized in Table 1.

**Table 1.** Clinical settings, sites, and indications for 32-gauge study needle use.

| Category | Physicians,<br>n | Needle uses,<br>n (%) |
| --- | --- | --- |
| Operator training level |  |  |
| Attending physician | 8 | 110 (100) |
| Clinical site |  |  |
| Byers Eye Institute, Stanford, CA, USA | — | 75 (68.2) |
| Erie Retina Research, Erie, PA, USA | — | 25 (22.7) |
| Taipei Veterans General Hospital, Taiwan | — | 10 (9.1) |
| Procedural setting |  |  |
| Outpatient clinic | — | 90 (81.8) |
| Operating room | — | 20 (18.2) |
| Clinical indication* |  |  |
| Ocular oncology | — | 56 (50.9) |
| General retina services | — | 51 (46.4) |
| Genetic retinal disease | — | 3 (2.7) |

\*Ocular oncology indications included uveal melanoma, vitreoretinal lymphoma, serial oncologic sampling, plaque brachytherapy, enucleation, and transscleral biopsy. Retinal disease indications included geographic atrophy, diabetic macular edema, and inherited retinal disease.

### Device Specifications

All study procedures used the STERiJECT Ophthalmic Low Dead Space (LDS) needle (TSK Laboratory, Tochigi, Japan), a 32-gauge needle with a 4-mm shaft designed for ophthalmic use and FDA-cleared. The device has a transparent, externally threaded, Luer-lock-compatible low-dead-space hub with a specified dead volume of 2.8 µL^22^ (Figure 1). Device specifications, with comparison to the standard 30-gauge hypodermic needles with a 12.7-mm shaft and fixed-needle 1-cc insulin syringes, are summarized in Table 2.

**Figure 1.**
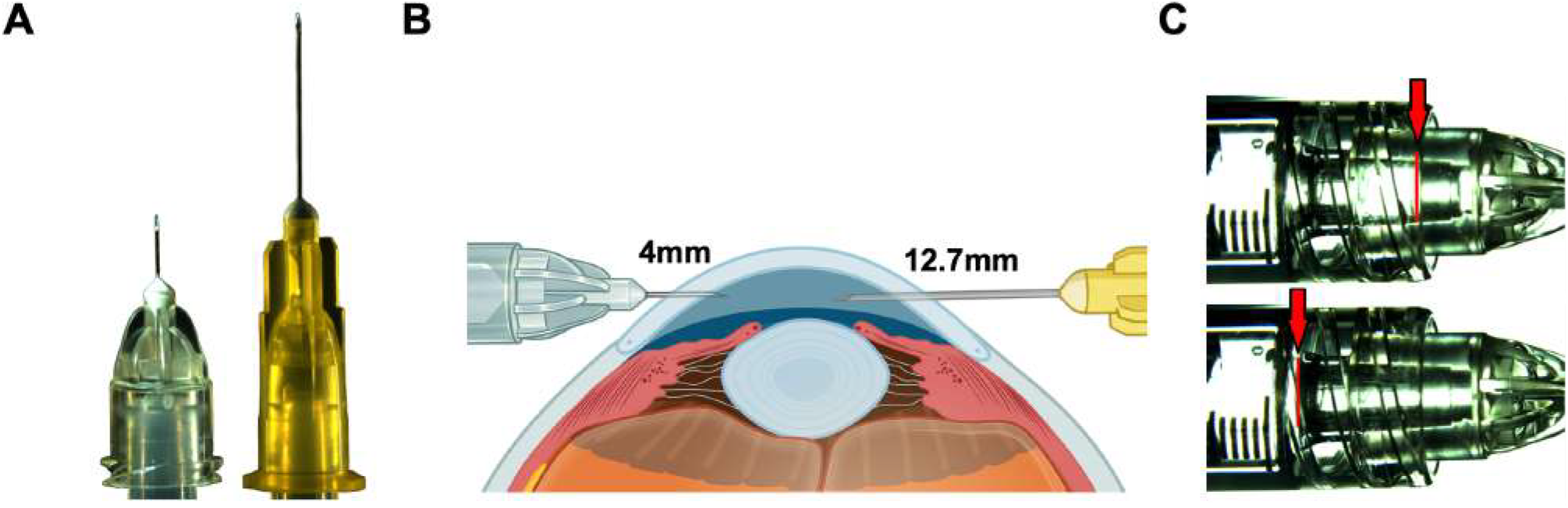
Comparison of the STERiJECT 32G *×* 4 mm ophthalmic needle and standard 30G hypodermic needle. **A**. Photographs comparing the STERiJECT ophthalmic needle (left) and standard 30G hypodermic needle (right). **B**. Schematic comparison of the 4-mm STERiJECT ophthalmic needle and the conventional 12.7-mm (½-inch) 30G hypodermic needle during anterior chamber paracentesis, illustrating the reduced intraocular needle length in the ophthalmic-specific design. **C**. Representative photographs of the transparent low-dead-space hub during aqueous humor aspiration. The advancing fluid meniscus is directly visible through the hub wall (red arrows), allowing the operator to confirm successful aqueous entry in real time without relying on plunger feedback. The upper and lower panels show the meniscus at sequential stages of aspiration; the fluid column fills the hub with minimal residual volume between the needle base and the syringe interface.

**Table 2.** Comparison of the 32-gauge ophthalmic needle with needles commonly used for anterior chamber paracentesis.

| Parameter | STERiJECT 32G × 4 mm ophthalmic needle | Standard 30G needle | Fixed-needle insulin syringe |
| --- | --- | --- | --- |
| Device category | Detachable ophthalmic low-dead-space needle | Detachable standard hypodermic needle | Integrated needle–syringe unit |
| Product identifier | VLDS-32004-050 | Product specific | Product specific |
| Gauge × length | 32G × 4 mm (5/32 in) | Typically 30G × 12.7 mm (1/2 in)** | Typically 29G–31G × 8–12.7 mm** |
| Outer diameter | 0.26 mm | Approximately 0.31 mm | Approximately 0.25–0.33 mm |
| Intended use | Ophthalmic use, including intravitreal use in the United States* | General hypodermic injection or aspiration | Subcutaneous insulin administration |
| Hub design | Detachable low-dead-space hub | Conventional detachable Luer hub | Fixed-needle design |
| Syringe compatibility | Attaches to a compatible syringe or male fitting | Attaches to a compatible Luer syringe | Fixed syringe; components cannot be interchanged |
| Dead space | 2.8 $\mu\text{L}$<sup>†</sup> | 6.80 ± 0.39 $\mu\text{L}$<sup>‡</sup> | 0.34 ± 0.28 $\mu\text{L}$<sup>§</sup> |
| Sterility and use | Gamma sterilized; sterile, single use; 5-year shelf life | Sterile and single use; manufacturer dependent | Sterile and single use; manufacturer dependent |
| Material considerations | Stainless-steel cannula; latex-free components | Manufacturer dependent | Manufacturer dependent |
| Ophthalmic-specific evaluation | Ocular, intraocular, and intracameral irritation testing; ophthalmic particulate and endotoxin standards referenced | Not designed or evaluated specifically for ophthalmic use | Not designed or evaluated specifically for ophthalmic use |
| AC tap-relevant design | Short 4-mm, 32G cannula with detachable low-dead-space hub | Familiar detachable needle, but longer 12.7-mm cannula | Low-dead-space integrated system, but dimensions and specifications vary |

\*United States labeling specifies ophthalmic use including intravitreal use; labeling in Japan, Canada, the European Union, and Australia also includes intracameral use. \*\*Specifications for standard 30G needles and insulin syringes vary by manufacturer and model.
Dead/residual volume values are based on device-specific or experimentally tested configurations. The STERiJECT Ophthalmic Needle Low Dead Space (LDS) has a specified dead volume of 2.8 $\mu\text{L}$ <sup>†</sup>,<sup>22</sup>. A residual volume of 6.80 ± 0.39 $\mu\text{L}$ <sup>‡</sup> was measured for a BD 30G × 8-mm needle paired with a BD 1-mL syringe,<sup>29</sup> and a residual volume of 0.34 ± 0.28 $\mu\text{L}$ <sup>§</sup> was measured for a BD Ultra-Fine fixed-needle syringe.<sup>30</sup> Residual volume varies by needle–syringe configuration.

### Procedural Settings and Surgical Technique

Procedures were performed in one of two clinical environments: the outpatient clinic or the ophthalmic operating room. In the outpatient clinic, patients were positioned at the slit-lamp biomicroscope or supine in a procedure chair, with the procedure performed under direct visualization in the latter case. Topical anesthesia was achieved with repeated instillations of proparacaine hydrochloride 0.5% or lidocaine hydrochloride 3.5% gel. Antisepsis consisted of povidone-iodine 5% applied to the conjunctival fornix and lid margins, followed by placement of a sterile lid speculum or manual lid retraction, according to physician preference. In the operating room, the tap was integrated into the sterile surgical workflow under operating-microscope visualization; patients received topical or monitored anesthesia care, and standard sterile draping was applied.

The paracentesis technique was standardized across settings. The needle was mounted on a 1-mL Luer-lock syringe with the plunger seal released before use. The needle was oriented parallel to the iris plane and introduced into the AC through peripheral clear cornea, just anterior to the limbus. Aqueous humor was aspirated slowly to a target volume of 50–100 µL. After needle withdrawal, a sterile cotton swab was applied to the entry site to confirm wound closure and exclude visible leakage. Specimens were transferred immediately to microcentrifuge tubes, placed on dry ice, and stored at −80°C in accordance with established ocular liquid-biopsy biobanking practices. Intraoperative events, including failure to enter the AC, were documented in the clinical record at the time of the procedure.

### Clinical Safety and Usability Endpoints

Each participating physician completed a standardized survey assessing four prespecified domains: (1) needle entry, handling, and positioning; (2) aspiration and sample recovery; (3) procedure-related adverse events; and (4) comparative performance relative to the standard 30-gauge needle. Items in domains 1 and 2 were rated on a 5-point ordinal scale (1 = poor, 2 = fair, 3 = neither agree nor disagree, 4 = good, 5 = excellent). Adverse-event items in domain 3 were recorded as binary (yes/no) responses, with a free-text field for other events. Comparative items in domain 4 assessed physician preference between the study needle and the standard 30-gauge needle, with response options favoring the 32-gauge study needle, favoring the 30-gauge needle, or no preference. The survey also captured physician characteristics, including years in practice, subspecialty, prior AC tap experience, number of AC taps performed with the study needle, and visualization method used for AC taps (direct visualization, slit lamp, or both depending on clinical setting), as well as average collected aqueous humor volume in the syringe, and device preference for a future AC tap.

Needle entry, handling, and positioning comprised five items: (a) smooth corneal entry with minimal resistance; (b) clean, crisp wound architecture without drag or tearing; (c) adequate needle control and maneuverability, including stability without excessive flexing or bending; (d) ease of achieving and maintaining desired AC position with minimal repositioning; and (e) adequate tip visibility while tracking in the AC. Aspiration and sample recovery comprised four items: (a) consistent, controlled aspiration flow with minimal resistance; (b) maintained flow without suspected occlusion or clogging; (c) minimal hub dead space; and (d) satisfactory sample volume recovery.

Adverse events comprised 12 items: lens touch; anterior capsule rupture; iris trauma, prolapse, or iridodialysis; hyphema or intraocular bleeding; AC shallowing or collapse during the tap; wound leak requiring suture or stromal hydration; persistent hypotony (intraocular pressure *<* 6 mm Hg at the end of the procedure); iatrogenic cataract; endophthalmitis within a 2-week follow-up window; retinal complication (tear or detachment); needle bending, breakage, or tip separation; unplanned conversion to a different needle or device.

Comparative performance relative to the standard 30-gauge needle was rated for five items: (a) corneal entry and wound integrity, (b) needle control and stability, (c) aspiration flow and resistance, (d) sample recovery, and (e) intra-procedure safety. Finally, each physician indicated the device they would choose for their next AC tap (study needle vs. 30-gauge needle, or no preference).

### Statistical Analysis

Descriptive statistics summarized procedure and physician characteristics. Categorical variables, including ordinal survey responses, were reported as counts and percentages. The primary safety outcome, defined as the proportion of procedures with any prespecified procedure-related adverse event, was reported with exact 95% confidence intervals using the Clopper–Pearson method. Safety outcomes were analyzed at the procedure level (n = 110), and survey outcomes at the physician level (n = 8).

## Results

### Study Cohort and Surgeon Characteristics

A total of 110 documented needle uses by eight physicians were included. Procedures were performed at three sites: an academic center, a community-based retina practice, and an international academic hospital, in outpatient clinic (90 uses, 81.8%) and operating-room (20 uses, 18.2%) settings. Clinical indications included ocular oncology (56 uses, 50.9%), general retina services (51 uses, 46.4%), and genetic retinal disease (3 uses, 2.7%). All eight retina specialists completed the survey spanning experience from 5 to more than 20 years in practice. Preferred technique varied: five performed taps with the patient supine under direct visualization, one at the slit lamp, and two alternated between approaches according to the clinical setting. Cohort characteristics are summarized in Table 1, and device specifications relevant to AC paracentesis are summarized in Table 2.

### Safety and Technical Outcomes

No ocular complications occurred among the 110 needle uses (0%; exact 95% CI, 0%–3.3%). There were no instances of lens touch; anterior capsule rupture; iris trauma, prolapse, or iridodialysis; hyphema or intraocular bleeding; AC shallowing or collapse, wound leak requiring suture or stromal hydration; persistent hypotony; iatrogenic cataract; endophthalmitis within the 2-week follow-up window; or retinal tear or detachment. No needle bent, broke, or tip separated during use, and no procedure required unplanned conversion to another needle or device. No additional events were reported in the free-text field. Two technical events were documented (1.8%; exact 95% CI, 0.2%– 6.4%). In both cases, needle entry remained partial thickness within the corneal stroma and did not reach the AC, resulting in failure to aspirate aqueous humor; the operator withdrew and repositioned the needle before successfully entering the AC. Neither event caused corneal injury or affected visual function over a six-month follow-up.

### Needle Entry, Handling, and Positioning

All five entry, handling, and positioning survey items were rated good or excellent on the 5-point scale from poor to excellent, with no ratings of poor, fair, or neither agree nor disagree (Figure 2A). Needle control and maneuverability and ease of maintaining AC position received uniformly excellent ratings (100%), smooth corneal entry with minimal resistance and adequate tip visibility were each rated excellent by most physicians (87.5%). Clean, crisp wound architecture without drag or tear-ing was rated excellent by the majority (62.5%), with the remainder rating it good (37.5%).

**Figure 2.**
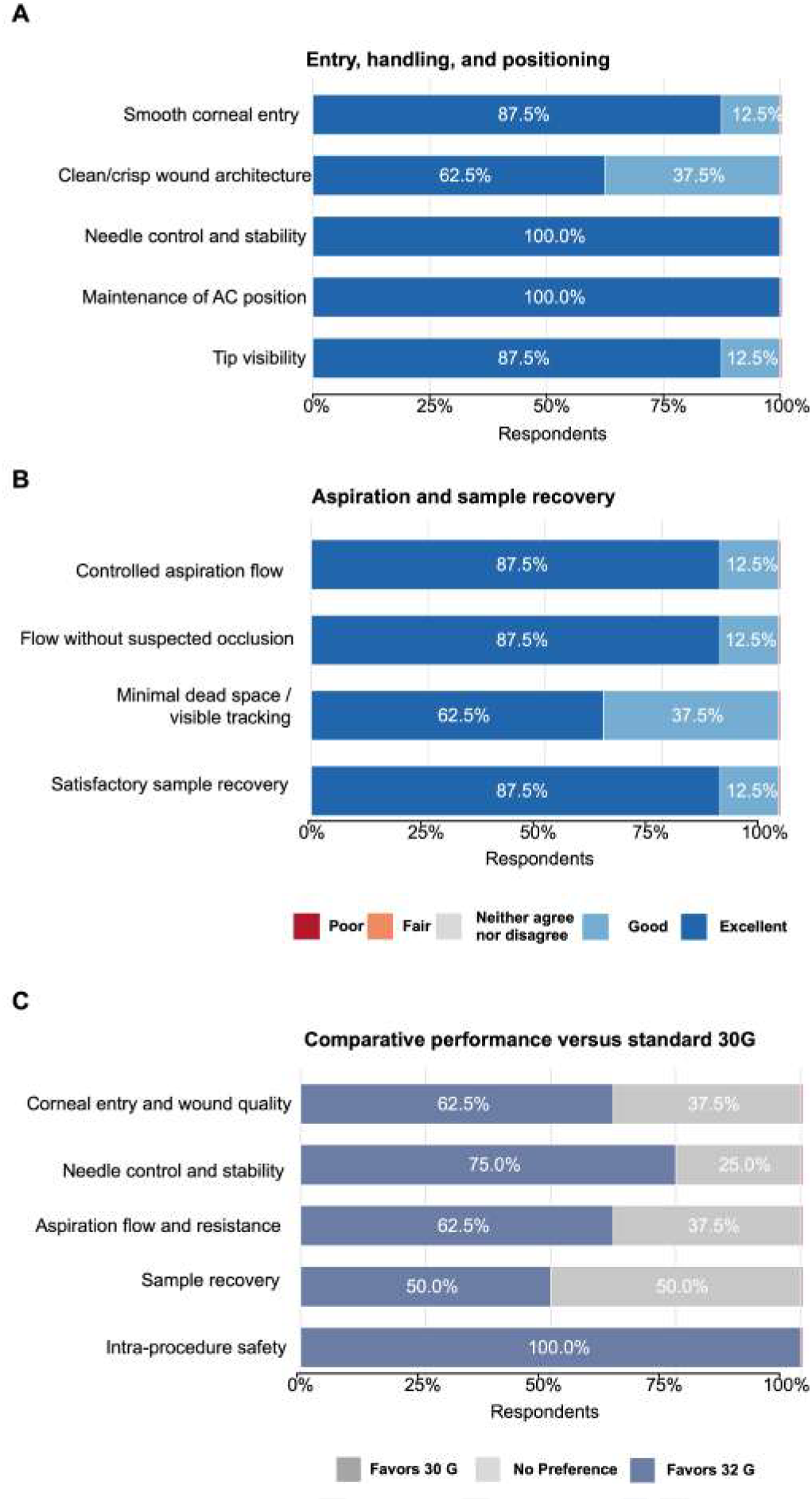
Physician assessment of the study needle and comparison with a standard 30-gauge needle. **A**. Ratings of needle entry, handling, and positioning on a 5-point scale from poor to excellent. **B**. Ratings of aspiration and sample recovery on the same scale. **C**. Comparative performance of the study needle versus a standard 30-gauge needle for corneal entry and wound quality, needle control and stability, aspiration flow and resistance, sample recovery, and intra-procedure safety.

### Aspiration and Aqueous Humor Sample Recovery

All four aspiration and sample-recovery survey items were likewise rated at least good or excellent. Consistent aspiration flow, maintained outflow without occlusion, and satisfactory collected sample volume were each rated excellent by most physicians (87.5%). Minimal hub dead space with visible sample tracking in the hub was rated excellent by the majority (62.5%), with the remainder rating each item good (37.5%) (Figure 2B). The average acquired aqueous humor volume in the syringe was 0.10 mL (range, 0.05–0.15 mL). These volumes reflect the protocol’s intentionally limited aspiration target of 50–100 µL rather than constrained recovery: comparable aqueous volumes support comprehensive downstream analysis, with more than 5,900 proteins quantifiable from as little as 50 µL of aqueous humor,^13^ and 50–100 µL specimens routinely sufficient for established multi-omic workflows.^1,8^

### Comparative Performance and Device Preference

Physicians rated the study needle relative to the standard 30-gauge needle on five comparative items, each on a 5-point scale from much worse to much better. The study needle was rated better or much better for intra-procedure safety by all physicians (100%), for needle control and stability by most (75.0%), for corneal entry and wound quality and for aspiration flow and resistance by the majority (62.5% each), and for sample recovery by half (50.0%). No item was rated worse or much worse. For the next AC tap, all physicians (100%) chose the study needle over the 30-gauge needle or no preference (Figure 2C).

## Discussion

In this multicenter study, we evaluated a short 32-gauge low-dead-space ophthalmic needle for AC paracentesis across academic, community, and international practice settings. No pre-specified adverse event occurred. All performance domains were rated good or excellent by all physicians, and no comparative aspect was rated worse than the standard 30-gauge needle. All surveyed physicians selected the study needle for their next AC tap. Although the safety of AC paracentesis has been established in retrospective series, the instruments themselves have never been the subject of structured, surgeon-reported evaluation. This study provides the first surgeon-reported assessment of a needle designed specifically for AC paracentesis, adding device-level evidence not captured in chart review.

Our safety findings align closely with prior series. A retrospective electronic health record analysis of 1,418 aqueous biopsies identified a single minor complication,^19^ and a multicenter study of 1,203 pediatric paracentesis procedures reported a similarly favorable profile.^20^ We observed no adverse events, consistent with the very low complication rates in these larger cohorts. Importantly, most procedures in our cohort were standalone taps performed in awake patients in the outpatient clinic, the setting least represented in existing safety data and the one where AC sampling for molecular applications will usually occur. Two technical events occurred, both partial-thickness corneal passes that did not reach the AC. Both resolved with withdrawal and repositioning, and neither left sequelae. We do not view this pattern as device failure. A short 4-mm shaft offers limited working length beyond the corneal tunnel, and a slightly shallow entry angle can leave the tip within the stroma. Both events were harmless because an intrastromal pass stops on its own, which is arguably a safer way to fail than the deep overshoot a longer 12.7-mm shaft allows. Attention to entry angle during early uses may reduce this learning-curve issue further.

The choice of a finer gauge needle is supported by the intravitreal injection literature, where needle downsizing has been studied beyond just AC sampling. Randomized comparisons show that finer needles cause less injection-site pain than larger bores,^23^ and 32-gauge needles have been associated with smaller postinjection rises in intraocular pressure without any loss of procedural performance.^24^ The traditional argument against finer needles has been mechanical: narrower shafts are more prone to bending and deformation during tissue entry, and the resulting loss of stiffness can compromise control. A short shaft counters this directly, and the effect can be quantified. For a cantilevered tube, tip deflection under load scales with the cube of shaft length but only with the fourth power of diameter through the cross-section’s moment of inertia;^25,26^ applying standard beam mechanics to the dimensions, the 4-mm 32-gauge shaft is approximately 16-fold more resistant to tip deflection than a standard 12.7-mm 30-gauge shaft, despite its finer gauge, because the 32-fold stiffness gain from the shorter length far outweighs the roughly 2-fold loss from the narrower cross-section. Consistent with this, no needle bent or broke in our study, and physicians rated needle control and stability better than with the 30-gauge standard. Our comparative ratings followed a nuanced pattern overall, which strengthens their credibility. Endorsement was strongest for intra-procedure safety, rated better or much better by every physician, and for needle control and stability, the domains most plausibly tied to the shortened shaft. Prior retrospective safety studies answer only the first question of an aqueous humor liquid-biopsy program, which is whether the eye is harmed.^19,20^ Chart review, however, does not address the questions that determine whether aqueous humor sampling can scale into multi-omic practice. It cannot tell us whether the specimen is recovered in analyzable volume, whether the instrument behaves predictably in the operator’s hands, or whether surgeons would adopt it over the repurposed needles they currently use.^21,27^ Complications get charted in retrospective studies, but specimen adequacy and operator experience do not. In absolute terms, recovery performed well: physicians rated aspiration flow, hub dead space, sample tracking, and recovered volume as good or excellent, and reported acquired volumes (average, 0.10 mL) at the upper end of the 50 to 100 µL protocol target. Whether recovery is better than with a 30-gauge needle is a separate question, and here surgeons were appropriately split, with half rating the device similar to the standard, perhaps because a narrower lumen draws fluid more slowly and may offset what the low-dead-space hub saves. These microliters are worth resolving, because more than 5,000 proteins and several thousand metabolites can be quantified from as little as 50 µL of well-preserved aqueous humor,^13^ and a conventional hub that retains tens of microliters can consume a meaningful fraction of such a specimen.

As aqueous humor liquid biopsy moves from targeted PCR to multi-omic profiling,^8,13^ the assays have become extraordinarily sensitive to specimen quality and volume, yet the step that determines both, collection itself, remains the least standardized part of the pipeline. A recent international survey found that clinicians perform AC taps with a wide variety of needles borrowed from hypodermic and intravitreal use,^21,27^ and lens touch with traumatic cataract has been reported with these standard instruments.^15^ The few studies that have tried to improve the procedure focused on the syringe, testing one-handed handles, plungerless injectors, and prefilled systems.^16–18^ A recent randomized trial compared a vacuum-based aqueous humor collector with an insulin syringe,^28^ but to our knowledge no study has evaluated the needle itself with physician-reported measures, even though the needle is what enters the eye and what determines wound size, working length, and dead space. Beyond the results for this particular ophthalmic needle, the survey structure used here, with prespecified safety items, ordinal ratings organized by domain, and comparison against the prevailing 30-gauge standard, can be reused to evaluate any AC sampling instrument. This standardized, instrument-level evidence is needed in multicenter trials and biobanking programs as aqueous humor sampling becomes part of the protocols.^1,11^ Finally, the short shaft may make this needle a good teaching instrument for residents and vitreoretinal fellows, because a 4-mm needle physically is less likely to reach the structures a trainee must avoid touching.

Several limitations should be addressed. Needle performance and comparative outcomes were physician-reported and unmasked, so social desirability and acquiescence bias cannot be excluded, although the graded pattern of comparative responses argues against uniform inflation. The 30-gauge comparison reflected recalled experience rather than a randomized within-surgeon design. Finally, we did not assess patient-reported outcomes, including comfort during awake in-clinic paracentesis, where the intravitreal literature suggests finer gauges may offer some benefit.^23^ Patient comfort will matter increasingly as liquid biopsy expands from single diagnostic taps to serial sampling for disease monitoring, where willingness to undergo repeat procedures becomes part of feasibility.

In conclusion, this study provides the first structured, multi-center evaluation of a needle designed specifically for AC paracentesis and supports its safe, controlled use for low-volume aqueous humor sampling. More importantly, it highlights specimen acquisition as a modifiable and standardizable component of the aqueous humor liquid biopsy workflow. As molecular profiling of aqueous humor expands across retinal diseases, purpose-designed collection devices may provide the procedural consistency needed for reproducible multicenter clinical trials, longitudinal sampling, and ultimately, clinical translation.

## Data Availability

All data produced in the present study are available upon reasonable request to the authors

## Credit Authorship Contribution Statement

VBM had full access to all the data in the study and takes responsibility for the integrity of the data and the accuracy of the data analysis. *Study concept and design:* VBM, AS, TCY. *Acquisition of data:* AS, TCY, KJ. *Surgical sample collection:* PM, CB, AM, JBL, SAS, SS, TCY, DRP. *Analysis and interpretation of data:* AS, TCY, PM, VBM. *Drafting of the manuscript:* AS, TCY, VBM. *Critical revision of the manuscript for important intellectual content:* all authors. *Statistical analysis:* AS, TCY. *Obtained funding:* PM, VBM. *Administrative, technical, and material support:* VBM. *Study supervision:* VBM.

## Institutional Review Board Statement

The study was approved by the Stanford University Institutional Review Board and adhered to the tenets set forth in the Declaration of Helsinki.

## Financial Support

VBM is supported by NIH grants R01EY039102, R01 EY037830 and P30EY026877 and by Research to Prevent Blindness, New York, New York. VBM and PM are supported by the Irene and Alan Adler Center for Ocular Oncology. TCY is supported by T32EY027816.

### Role of the Sponsor

The funding organizations had no role in design and conduct of the study; collection, management, analysis, and interpretation of the data; preparation, review, or approval of the manuscript; and decision to submit the manuscript for publication.

## Notes

### Competing Interest Statement

The authors have declared no competing interest.

### Author Declarations

Stanford University Institutional Review Board of Stanford University gave ethical approval for this work (IRB-85576).

